# O Group is a protective factor for COVID19 in Basque population

**DOI:** 10.1101/2020.07.13.20152447

**Authors:** M Muñoz-Culla, A Roncancio-Clavijo, B Martínez, M Gorostidi, L Piñeiro, A Azkune, A Alberro, J Monge-Ruiz, T Castillo-Trivino, A Prada, D Otaegui

## Abstract

ABO blood groups have been related to COVID19 infection. ABO blood groups are slightly differently distributed in the populations and therefore these results should be replicated in the specific areas with a proper control population. In this work, we present data from 412 COVID19 patients and 17796 blood donors from Gipuzkoa, a region in Northern Spain. Our data shows the importance of group O as a protective factor.

## Introduction

COVID19 is a pandemic disease caused by Severe Acute Respiratory Syndrome Coronavirus 2(SARS-CoV-2). This disease has rapidly become the most important world Health challenge in the last century.

Despite an incredible and collaborative research effort in the last months, the pathogenesis and the clinical symptoms of COVID19 are poorly understood. Several works have been published with the aim of understanding the great heterogeneity in the infection and the clinical manifestations of SARS-CoV-2 in different patients. Several biomarkers have been proposed as risk or protective factors. In this scenario, a recent paper, demonstrating the importance of collaborative networks, presents the results of a Genomewide analysis focused on COVID19 patients (1). One of the associated locus in chromosome 9, point to an association between ABO blood group and risk of infection by SARS-CoV-2. This association with the ABO blood group has been previously reported in other series showing that Group A is a risk factor while Group O seems to be a protective factor for SARS-CoV-2 infection.

ABO group is differentially distributed according to the ethnic origin and therefore the results from these studies are highly dependent on the distribution of the control group in each region. With this in mind, our objective in this short report is to study the ABO group distribution in COVID19 patients in Gipuzkoa (Basque country).

## Methods

Blood group from COVID19 patients were retrieved from electronic clinical history. All the included patients had tested positive for SARS-CoV-2 infection by PCR at the Microbiology Department from Donostia University Hospital.

The distribution of the ABO group from COVID19 patients was compared to that from the general Basque population, obtained from the database of the Basque Blood Bank, preserving the anonymization of all the participants.

Data were analyzed with SPSS (v 20). Distribution was analyzed using chi-square test.

Mild cases were defined as having mild clinical symptoms with no signs of pneumonia on imaging. Moderate cases were those individuals showing fever and respiratory symptoms with radiological findings of pneumonia, and severe cases were those meeting any of the following criteria: (1) Respiratory distress (≧30 breaths/ min); (2) Oxygen saturation≤93% at rest; (3) Arterial partial pressure of oxygen (PaO2)/ fraction of inspired oxygen (FiO2)≦300mmHg (l mmHg=0.133kPa).

## Results

Our cohort includes 412 COVID19 patients and 17796 anonymous blood donors from the same geographical area (Gipuzkoa). Average age of COVID patients is 57.64 yrs, with a 68.45 % of women. 49.29 % of the classified patients shows moderate clinical symtomps, 8.92 % mild and 41.78 % severe.

The distribution of the ABO groups is shown in Table 1.

**Table 1:** ABO group distribution in the Gipuzkoa cohort. P-values from the Chi-square test are shown for the comparison of each group distribution in blood donors vs COVID19 patients.

|  | A | B | AB | O | TOTAL |
| --- | --- | --- | --- | --- | --- |
| **Blood donors** | **40.65% (7235)** | **5.16% (918)** | **1.20% (355)** | **52.19% (9288)** | **17796** |
| **Men** | 40.59% (4135) | 5.38% (548) | 2.05% (209) | 51.98% (5296) | 10188 |
| **Women** | 40.75% (3100) | 4.86% (370) | 1.92% (146) | 52.47% (3992) | 7608 |
| **COVID19 patients** | **48.30% (199)** | **8.74% (36)** | **3.88% (16)** | **39.08% (161)** | **412** |
| **Men** | 53.85% (70) | 6.92% (9) | 2.31% (3) | 36.92% (48) | 130 |
| **Women** | 45.75% (129) | 9.57% (27) | 4.61% (13) | 40.07% (113) | 282 |
| **p-value** | 0.00179 | 0.00186 | 0.012204 | <0.0001 |  |

According with the literature, in our data group A is more frequent in COVID19 patients (48.3 % vs 40.65 %, p= 0.00179) while group O is less represented in this group (39.08 vs 52.19 %, p<0.0001). The group A presents an OR of 1.36 (CI 95% 1.12-1.66, p=0.0019) while group O present an OR of 0.5876 (CI95% 0.481-0.7177, p<0.0001). If ABO group is analyzed as a dichotomic variable; defined as having any antigen (A,B and AB) or none (O), the results highlight the protective role of group O, the OR for noO is 1.7019 (CI95% 1.94-2.08, p<0.0001)

Although the ABO group distribution is similar between women and men in the control group, it presents differences in the COVID19 group. This different distribution by sex becomes relevant in the two affected groups (A and O) that seem to indicate a higher risk for infection in men.

No difference has been found in ABO group distribution with severity.

## Discussion

Our data shows the importance of the absence of immune antigens defined by group O as a protective factor for Sars-CoV-2 virus infection. These results are consistent with the preprint works conducted in the Chinese population(2) and in the New York cohort (3), and also with the genotype observation in the Italian and Spanish population (including samples from Gipuzkoa). Due to the different distribution of the ABO groups by ethnicity the validation of these kind of observations in each region and with the proper controls is important to achieve a better understanding of how the virus is spreading in the population. In our study, the group O distribution differs from that of other series seeming more protective in Gipuzkoa population (fig 1).

**Fig 1:**
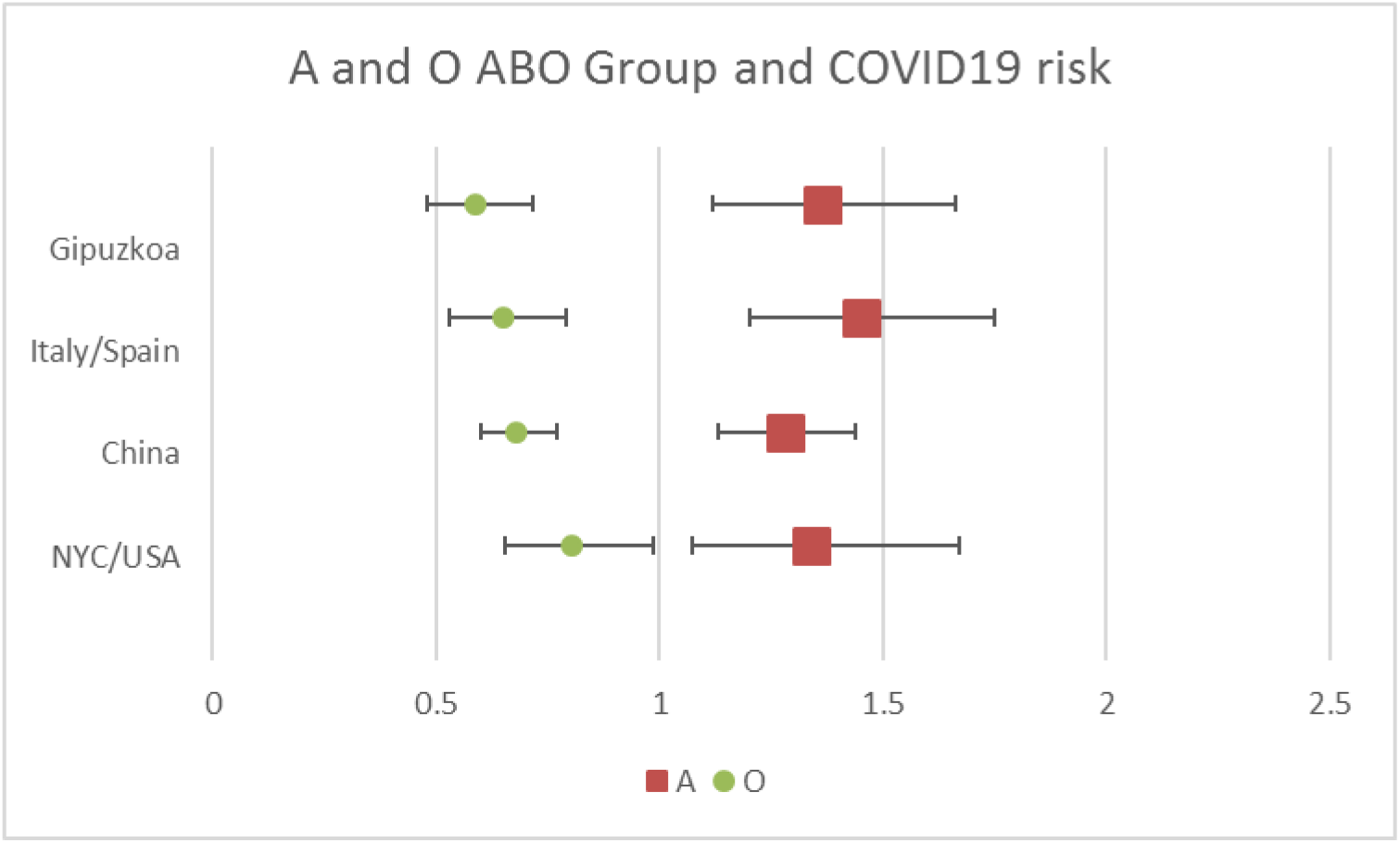
Odd ratio of A (red square) and O group (green circle) from our study and the ones found in the literature.

The observed different distribution seems to be related to the infection event and not with the course of the disease. The mechanisms behind these observations remain unknown. It has been proposed that the presence of anti-A Antibodies could be protective against viral entry into lung epithelium or it may be the fact that O group presents an altered glycosiltransferase activity and therefore, an increased clearance of Von Willebrand factor, that could protect O group patients from the COVI19-related microvascular thrombosis and endothelial dysfunction(4).

In conclusion, our data confirm the idea that O group is a protective factor for COVID19 but also supports the need for further studies in each region with proper population controls. The biological implications of these observations deserve future investigations to shed light on the mechanisms behind COVID19 infection risk.

## Data Availability

All the referred data of the article are available

## ACKNOWLEDGMENTS

Authors want to thank all the clinicians that have work so hard these last months and to all the COVID patients.

